# Surveillance of interhospital VRE transmission using routine centralised, multicentre whole genome sequencing

**DOI:** 10.1101/2025.10.02.25337224

**Authors:** Djamayl L.H.H. Engelen, Jean-Luc Murk, Veronica A.T.C. Weterings, Heike Schmitt, Bram M.W. Diederen, Andreas L.E. van Arkel, Jaco J. Verweij, Wouter van den Bijllaardt, Jeroen H.T. Tjhie, Joep J.J.M. Stohr

**Author notes:** Corresponding author is Djamayl L.H.H. Engelen, Microvida, Amphia Hospital, Molengracht 21, 4818 CK Breda, The Netherlands. **Author Note** Part of our data was previously presented at the ESCMID global 2024 congress in Barcalona during an oral presentation.

## Abstract

**Objective:** Whole genome sequencing (WGS) is increasingly being used to guide infection prevention and control measures. However, the value of regional prospective WGS surveillance for vancomycin-resistant *Enterococcus faecium* (VREfm) remains uncertain.

The objective of this study is to investigate the value of routine centralised multicentre WGS in regional outbreak detection and analysis of transmission of VREfm within and between healthcare facilities.

**Methods:** VREfm isolates identified from patient samples in five different hospitals in the Netherlands during a one-year period were whole genome sequenced and typed using core genome multilocus sequence typing (cgMLST). Sequence data were correlated with admission data of the year prior to the first positive VREfm culture of the respective patients. By combining epidemiological and sequence data within and between hospital transmission events were identified.

**Results:** 57 VREfm isolates were detected during the study period. 38/57 (66.7%) isolates clustered with at least one other isolate. Based on our definitions, intrahospital transmission could be demonstrated in 37 of 38 cases and interhospital transmission in 3 of 38 cases. Using this approach, a multicenter outbreak and two local outbreaks were detected.

**Conclusions:** Routine whole genome sequencing of VRE is a powerful tool for detecting, tracing and delineating outbreaks. When integrated into a centralised surveillance system spanning collaborative healthcare networks, it becomes an essential tool for unravelling the complex transmission dynamics of VRE within and between hospitals, ultimately strengthening infection control strategies.

## Introduction

Enterococci are one of the top three micro-organisms reported in hospital-acquired infections (1). Vancomycin-resistant enterococci (VRE) are associated with increased morbidity and mortality compared to infections with vancomycin-susceptible enterococci (2–5). In the Netherlands, the overall prevalence of vancomycin-resistant *Enterococcus faecium* (VREfm) colonization is unknown but studies performed in high-risk hospital wards report a prevalence of 1-8.6% (6,7). Furthermore, the prevalence of VREfm isolates in blood cultures is low compared to the European Union/European Economic Area (EU/EEA), possibly reflecting low colonization prevalence (8,9). Despite this, nosocomial outbreaks occur regularly with 137 VREfm outbreaks between 2012 and 2023 which accounts for 20-32% of all nosocomial outbreaks in the Netherlands. In 2021 and 2022 a total of nine outbreaks per year were reported (8). To prevent nosocomial VRE transmission, guidelines suggest infection prevention and control measures, including contact tracings with positive screening cultures for high-risk contacts (10). Whole genome sequencing (WGS) is increasingly used to guide infection prevention and control (11,12). Recently, the routine application of WGS for all multi-drug-resistant organisms (MDRO) isolates in the intrahospital surveillance showed that VREfm transmission could be detected at an earlier stage than by retrospective sequencing triggered by epidemiological data (11). However, the added value of routine WGS surveillance of VREfm at a regional, interhospital level is still uncertain. The present study aimed to evaluate the use of regional routine WGS of VREfm isolates in five hospitals across two provinces in the Netherlands to detect VREfm transmission within and between hospitals.

## Methods

### Setting

This prospective cohort study was performed from January 1, 2022, to August 1, 2023, across five hospitals (Elisabeth-TweeSteden Hospital in Tilburg, Amphia Hospital in Breda, Bravis Hospital in Roosendaal, Admiraal de Ruyter Hospital in Goes, Zorgsaam Hospital in Terneuzen), with a catchment area of 1.7 million people in the provinces North-Brabant and Zeeland in the Netherlands. The Elisabeth-Tweesteden Hospital is the reference centre for all complex trauma, neurological and neurosurgical patients in North Brabant.

In all five hospitals, patients are screened for VREfm when they meet criteria for screening based on national guidelines (**Table 1**)(10). New unexpected VREfm carriage or infection can also be detected in routine clinical cultures. Whenever a new VREfm was detected in any of the hospitals (routine clinical cultures, screening cultures and contact tracing cultures), the isolate was sent to the Elisabeth-Tweesteden Hospital for sequencing and cluster analysis.

**Table 1:** Criteria for VREfm screening based on national guidelines.

|  |
| --- |
| Patients admitted more than 24 hours to a foreign hospital < 2 months before admittance |
| Patients admitted to a foreign hospital 2-12 months before admittance and underwent an invasive procedure in the foreign hospital |
| Patients transferred from a hospital with an active VREfm outbreak |
| Patients living in an asylum seekers' centre |

Patients colonized or infected by VREfm, are admitted to single patient rooms and nursed using contact precautions. The VREfm colonization status is communicated between infection control departments of the different hospitals when patients are transferred between different hospitals. Contact tracings are performed when VREfm is detected in samples of patients that were not previously admitted to a single patient room and were not nursed using contact precautions prior to VREfm detection. Additionally, after cleaning and disinfection, VREfm environmental cultures of the room in which VREfm positive patients were admitted are performed. Only after negative cultures, the room is given free for the admission of new patients. When cultures show growth of VREfm, a new round of cleaning, disinfection and culturing is performed until negative cultures.

### Included patients

All patients (in- and outpatient) in whom a VREfm was detected in a sample sent for microbiological culture between January 1, 2022 and August 1, 2023, were included in the study. For each included patient, the admission history (admission and discharge dates, wards of admission and rooms of admission) of the past 365 days in all five hospitals was retrieved. Sample metadata (collection date, hospital where collection was performed, sampling site and patient location during sampling) from VREfm-positive samples were also registered.

### Detection of VREfm

When *E. faecium* were identified in samples using MALDI-TOF MS (Bruker Daltonics GmbH (Bremen, Germany)), susceptibility testing was performed using the BD Phoenix system (BD, Franklin Lakes, New jersey, US) or Vitek-2 (bioMérieux, Marcy-l’Étoile, France) depending on the hospital. For samples collected from sterile sites (e.g. blood cultures, cerebrospinal fluid), a confirmation test with a dose of 5 microgram vancomycin disk was used for disk diffusion following EUCAST guidelines (13). When reduced susceptibility to vancomycin was detected (MIC>4 mg/L, zone<12 mm or fuzzy border for disk diffusion) according to EUCAST breakpoints), molecular confirmation of VREfm was performed using GeneXpert using the Xpert *vanA*/*vanB* kit (Cepheid, California, USA) following the manufacturer’s instructions or using an in-house real-time PCR detecting the *vanA* or *vanB* gene as previously described (14).

### Molecular typing and cluster analysis

Whole genome sequencing was performed for the first VREfm isolate of each included patient using Nextera XT chemistry on a MiSeq sequencer (Illumina, San Diego, CA, USA) and assembled using CLC Genomic Workbench (Qiagen, Venlo, Netherlands). The following quality control criteria were used: coverage ≥20; number of scaffolds ≤1000; N50≥15,000 bases; and maximum scaffold length ≥50,000 bases. Species-specific core-genome multi-locus sequence typing (cgMLST) was performed for all sequenced isolates using Ridom SeqSphere Version 5.1.0 (Ridom, Münster, Germany). All-to-all pairwise genetic differences were calculated between the isolates by counting the total number of allelic differences in the cgMLST typing scheme. Clonality was defined as ≤20 allelic differences. Isolates with ≤ 20 allelic differences were assigned an identical local cluster type number (15). Time from detection of VREfm to the results from cluster analysis took around 2-3 days, and reports were sent directly to infection control teams.

### Data analysis

Possible transmission events were investigated using admission and sequence data for each new VREfm-positive patient. For each cluster type, the epidemiological relationship between the patients was evaluated with a look-back period of 1 year before the first positive VREfm culture. Intra- and interhospital transmission was investigated for clonal isolates and definitions are given in **Table 2**.

**Table 2:**
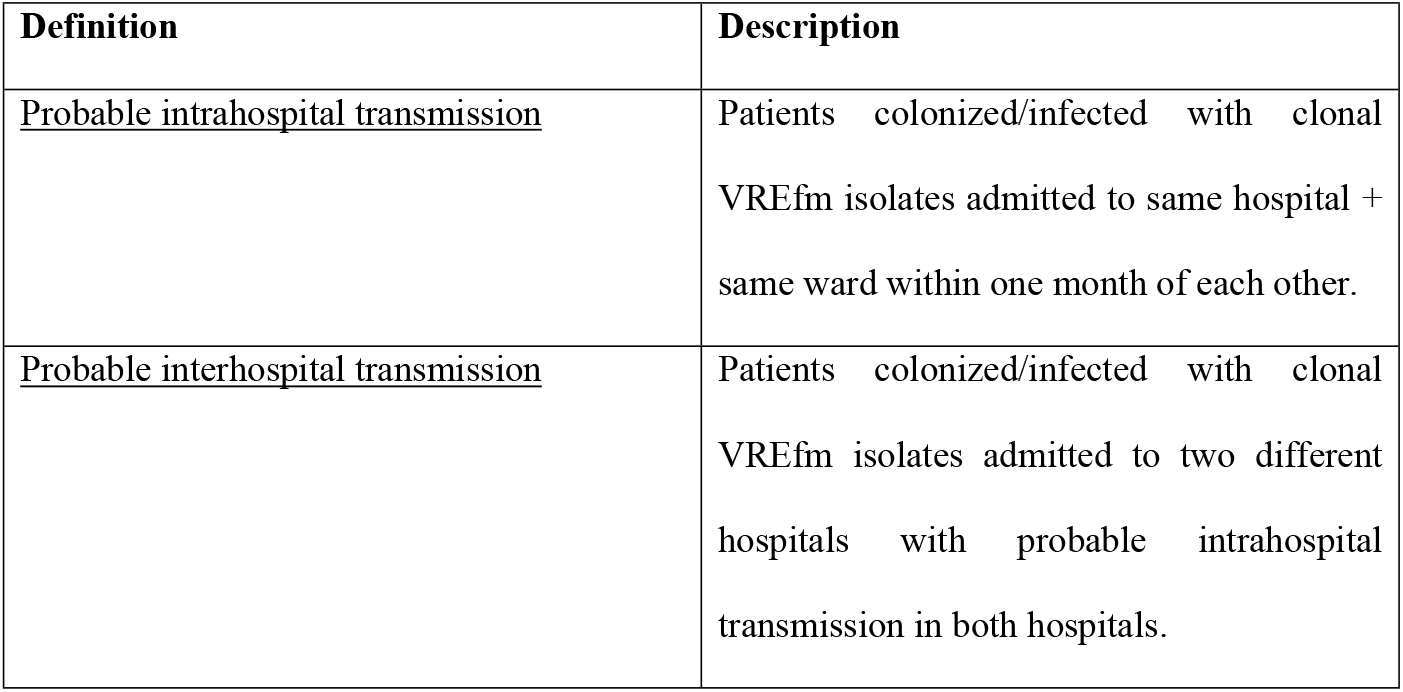
Definitions of transmission events.

### Ethics

The study was evaluated by the regional medical ethics committee METC Brabant (NW2023- 35) and did not fall under the scope of the WMO. Consent of individual patients was therefore not necessary

### Results

In the study period, 57 patients had a newly detected VREfm (**Table S1**). In one patient, VRE was isolated in a scrotal wound sample, in one in a blood culture, in 54 in rectal/perineal swabs, and in one in a stool sample. Additionally, in two patients VRE was also isolated in a urine sample. None of the identified VREfm-positive patients were previously known to be infected or colonized with VREfm. Forty-three patients were admitted to only one hospital in the year before their first positive VREfm culture, four patients were admitted to two different hospitals, and 10 were never admitted to a hospital in the year prior to their first positive culture (**Table S1**). The latter patients were sampled outside the hospital (n=4 in a rehabilitation center and n=1 by a primary care physician), in an outpatient clinic (n=2), or were screened upon admission (and placed in contact isolation while awaiting culture results) because they were previously admitted to a hospital in another country but were not admitted to any of the included hospitals in the year prior to the screening (n=3).

In 52 isolates, a *vanA* gene was detected and in 5 isolates a *vanB* gene. For 51 isolates a sequence type (ST) could be assigned amounting to 7 different STs. Thirty-eight of 57 VREfm isolates (66.7% of all patients, 84.4% of admitted patients) were clonally related to at least one other isolate in the study. Three different clusters of clonally related isolates were identified: cluster C1 (n=26 isolates); cluster C2 (n=6 isolates), and cluster C3 (n=6 isolates). Isolate-pairs of the same cluster had a median genetic distance of one allele (13 alleles within the 95^th^). Isolate-pairs not belonging to the same cluster had a median genetic distance of 367 alleles (444 alleles within the 95^th^ percentile) (**Figure S2**).

The isolates belonging to cluster C1 were detected in 26 patients admitted to three different hospitals: six patients were admitted to Hospital A, 16 to Hospital C, and four were admitted to two different hospitals (three to Hospital B and C; and one to Hospital A and C) (**Fig1**). Epidemiological analysis based on previously defined definitions (**Table 2)** revealed probable intrahospital transmission events involving 19 patients across six wards in hospital C: intensive care (IC), orthopaedic surgery, trauma surgery, urology/gynaecology, vascular surgery, and neurosurgery with an intricate transmission network between these different wards (**Fig2**). Patients with isolates belonging to cluster C1 admitted to hospital A (n=7) and hospital B (n=2), were all admitted to the neurology ward of their respective hospital, indicating probable intrahospital transmission events in both hospitals A and B (**Fig2; Table S1**). Additionally, probable interhospital transmission was identified in three out of four patients who were admitted to two different hospitals: These three patients were admitted to the neurosurgery or IC ward of hospital C and the neurology ward of either hospital A or B. (**Fig1; Fig2; Table S1**). The first positive VRE culture of those patients was taken in hospital A (VRE isolate 16) and hospital B (VRE isolate 19 and 23). Cluster C1 isolates were detected from February 2, 2022 to November 3, 2022 (**Fig1**). During this period, VREfm isolates not belonging to cluster C1 were also detected in six other patients admitted to hospital C. These isolates formed a separate cluster, Cluster C2. (**Fig1; Table S1)**. All patients with isolates belonging to cluster C2 were admitted to the geriatric ward in hospital C within one month of each other, pointing to probable intrahospital transmission. None of these patients were admitted to other hospitals (**Fig1; Table S1**). Cluster C3 involved isolates belonging to six patients, all of whom were admitted to hospital D. None of these patients were admitted to other hospitals (**Fig1**). A probable intrahospital transmission event of a clonal VREfm isolate with at least one other patient was identified in all 6 of the patients of cluster C3 (**Table S1**).

**Fig 1:**
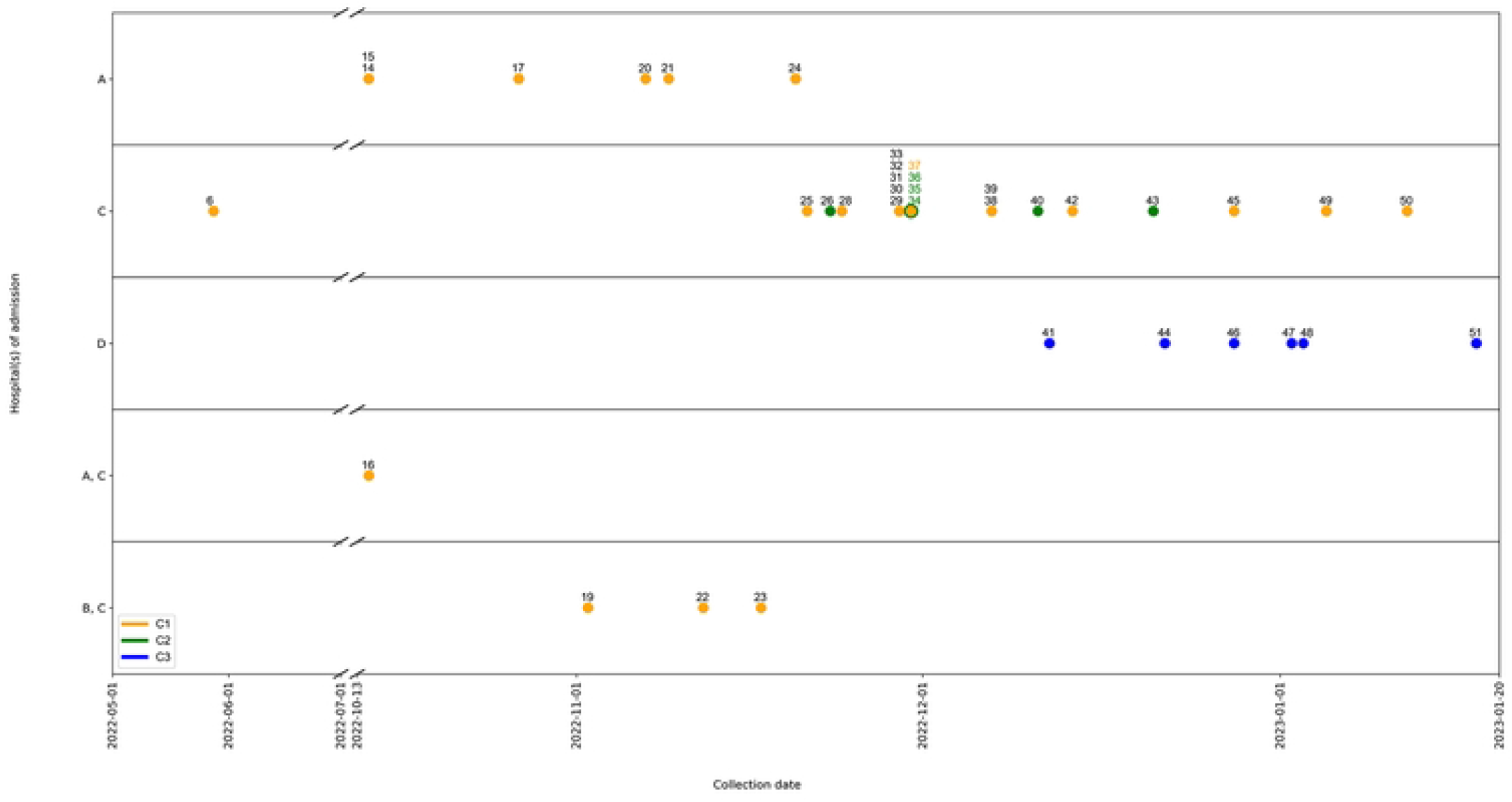
Admission data of VREfm positive patients of which their isolate clustered with at least another VREfm isolate. Numbers correspond to the respective VREfm isolates in Table S1. * Green numbers correspond to cluster C2. Yellow numbers to cluster C1.

**Fig 2:**
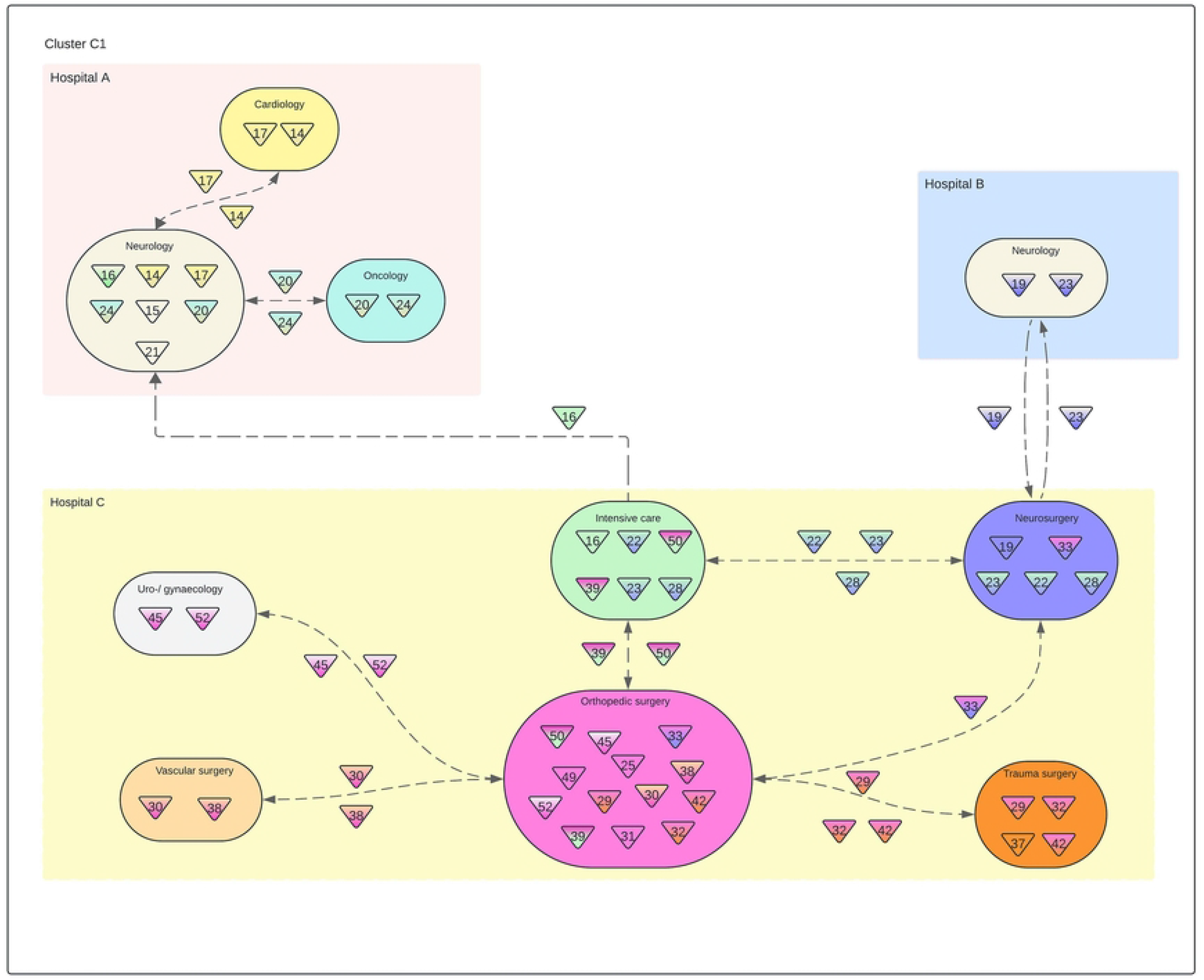
Patients with isolates belonging to cluster C1 and their wards of admission in hospital A, B and C. Only patients with a probable nosocomial transmission link are shown. Numbers in triangles: unique patient number; color of triangle: ward(s) of admission of patient corresponding tot the color of the respective wards.

## Discussion

The present study demonstrates that collaborative healthcare networks give rise to intricate VREfm transmission networks, which can be detected, traced and delineated by whole genome sequencing of VREfm. When integrated into a centralised surveillance system spanning collaborative healthcare networks, it becomes an valuable tool for unravelling the complex transmission dynamics of VRE within and between hospitals and other healthcare institutions, ultimately strengthening infection control strategies.

As whole genome sequencing is increasingly used to support infection prevention and control, its role is more and more established at the level of the individual hospital (11,12,16–18) The results of the present study show that the transfer of patients between hospitals can lead to interhospital transmission of VREfm, which is in line with other studies showing that collaborative healthcare networks with patient transfers are a risk factor for MDRO transmission and outbreaks within those networks (19). Extensive VRE transmission can occur before being noticed due to VRE’s propensity to spread and colonise asymptomatically and its ability to persist op surfaces for extended periods of time (ranging from days to years) (20). This may complicate the identification of epidemiological relatedness of VRE carriers, potentially delaying outbreak detection, particularly in cases of interhospital transmission. Routine WGS across collaborative healthcare networks has the potential to fill this gap of uncertainty by extrapolating its value at the level of the individual hospital. However there is still a paucity of data to support this at a multi-hospital level. Several other studies used multi- hospital prospective WGS to detect clusters spanning multiple hospitals, but these lacked a detailed discussion of the correlation between WGS data and epidemiological links at an interhospital level (11,17). Without sequence data every newly detected VREfm-carrying patient can be regarded as an isolated case or as part of ongoing transmission, depending on the spatio-temporal relationship with other VREfm-positive patients. Additionally, without a centralised approach, the detection of interhospital spread within collaborative healthcare networks is dependent on the communication between infection control teams of the respective hospitals which is often event-driven (e.g. around patient transfers), retrospective and/or fragmented. Furthermore, even if typing data is available in every involved hospital, it should be standardised in order to be correlated between the hospitals within the network. The present study shows that the addition of sequence data supported in delineating outbreaks (e.g. the discrimination of the C1 from the C2 cluster) and combined with a centralised approach also revealed interhospital transmission events which, combined with epidemiological data, could eventually be reconstructed into a large regional VRE outbreak. Due to the centralised nature of the data, it was possible to create a clear overview of the transmission network within and between the involved hospitals. Our results therefore contribute to the growing body of evidence for the value of routine centralised WGS within collaborative healthcare networks.

The current study has several limitations. Firstly, the relatively low number of VRE-positive patients detected during the study period outside the context of outbreaks and/or contact tracings introduced selection bias to our results that 86.4% of VREfm isolated from admitted patients were part of a genetic cluster. However, the finding that most VREfm isolates within hospitals are genetically related and indicate within-hospital transmission is in line with studies where clinical isolates were reported separately to reduce selection bias by contact investigations (11,21,22). An additional limitation is that the discrimination between intrahospital and interhospital transmission can be difficult based on the used criteria. For instance, when the epidemiological analysis of patients with clustering isolates reveals an epidemiological link between the patients in two or more hospitals (e.g. isolates 19 and 23) it will be difficult to determine in which hospital the transmission took place and therefore whether it should be classified as intrahospital transmission in hospital B or C or as interhospital transmission between both hospitals. However, in the interhospital transmission described between patients in hospital A and C with isolates belonging cluster C1 the possibility of intrahospital transmission could be ruled out.

Further studies should evaluate cost-effectiveness and whether our findings apply to other multidrug-resistant organisms.

## Conclusions

Collaborative healthcare networks go hand in hand with intricate transmission networks of VREfm and plead for regional infection prevention and control strategies. Routine, centralised WGS within those networks can play an important role as part of those regional strategies for the detection, tracing and delineation of transmission of VREfm.

## Data Availability

All whole genome sequencing reads-files are available from the ENA database (study accession number PRJEB96955) Sample metadata (sample origin, collection date) and anonymized epidemiological data of patients (hospital(s) and ward(s) of admission) is provided in the Supplementary Data. Exact date/time and hospital/ward-mutations of all patients during their admission are not included as we are of the opinion that this is too privacy sensitive and could be traced to individuals.

https://www.ebi.ac.uk/ena/browser/view/PRJEB96955

## Acknowledgements

We thank the staff of the microbiology department for storing and culturing all isolates. We also thank the members of the infection control unit for their role in outbreak management.

## Supporting Information

**S1 Table: Metadata of all VRE positive patients of the year previous to first positive culture per patient**. N/A: not admitted.

**S2 Fig: Distribution of pairwise genetic distance (alleles) between isolates within and outside a cluster**

**S3 Table: Distribution and relation of ST-type and cluster type**

## Notes

### Competing Interest Statement

The authors have declared no competing interest.

### Funding Statement

The author(s) received no specific funding for this work.

### Author Declarations

The study was approved by the regional medical ethics committee METC Brabant (NW2023-35).

